# Patterns of retention in care during clients’ first 12 months after HIV treatment initiation in South Africa: a retrospective cohort analysis using routinely collected data

**DOI:** 10.1101/2023.06.13.23291348

**Authors:** Mhairi Maskew, Mariet Benade, Amy Huber, Sophie Pascoe, Linda Sande, Sydney Rosen

**Affiliations:** Health Economics and Epidemiology Research Office, Faculty of Health Sciences, University of the Witwatersrand, Johannesburg, South Africa; Boston University School of Public Health, Boston, MA, USA

**Keywords:** South Africa, antiretroviral therapy, engagement, patterns, Tier.Net

## Abstract

**Background:** Retention in HIV care during the early treatment period is one of the most serious challenges facing HIV programs, but the timing and patterns of early disengagement from care remain poorly understood. We describe patterns of engagement in HIV care during the first and second 6-month periods after initiation.

**Methods:** We analysed retrospective datasets of routinely collected EMR data from ≥18-year-old clients who initiated ART at public sector clinics in South Africa after 01/01/2018 and had ≥14 months potential follow up. Using scheduled visit dates, we classified observed visits into “as planned” or “late” and characterized engagement in care as continuous (all scheduled visits attended ≤28 days late), cyclical (at least one visit >28 days late with a return visit observed) or disengaged (visit not attended and no return visit to the same facility observed).

**Results:** 33,821 client records were included (65% female, median age 33). By six months after ART initiation, 57% had remained continuously in care, 14% had engaged cyclically, 11% had transferred to another facility, 1% had died and 16% had disengaged from care at the initiating facility. Among disengagers in the first 6 months, 58% did not return after their initiation visit, 10% disengaged within the first three months, and the remaining 32% disengaged between 3-6 months. By 12 months after initiation, the overall proportion disengaged was 23%, and only 38% of the cohort had maintained continuous engagement for the full 12 -month period.. Patterns of engagement that were established during the first 6 months on ART demonstrated little change in months 7-12, with participants who were cyclically engaged in months 0-6 were nearly twice as likely to disengage in months 7-12 as continuous engagers in months 0-6 (relative risk 1.8, 95% CI: 1.70-1.99).

**Conclusions:** As recently as 2018, fewer than 60% and 45% of clients starting ART in South Africa were continuously engaged in care (no interruptions >28 days) at 6 and 12 months, respectively, at their initiating facilities. The needs of continuous and cyclical engagers and disengagers during the first 6 months after initiation may differ and require different interventions or models of care.

## Introduction

As high HIV-prevalence countries in sub-Saharan have neared global targets for HIV testing and for viral suppression among those remaining on treatment, the main challenge many countries face is retaining clients on antiretroviral therapy after treatment initiation. Retention on ART during a client’s first year after initiation, and in particular during the first six months, is especially problematic [1]. Two recent studies, one observational[2] and one a trial of a case management intervention[3], both estimated analysis estimated 6-month attrition from care at 26% in South Africa.

Although there is a large body of literature and multiple systematic reviews reporting rates of retention and reasons for attrition from HIV treatment[4–9], the precise timing and patterns of client disengagement from care are poorly described. To address this problem, several previous studies have described standard care “trajectories” among ART clients[10–12]. These studies have largely reported retention and attrition in multi-year aggregate time periods, however, masking the details of client behavior during the two critical early periods, 0-6 and 7-12 months after initiation.

In this study, we used retrospective medical record data from three provinces in South Africa to analyze patterns of care between ART initiation, 6 months after initiation, and 12 months after initiation. Results show exactly when during these periods clients are disengaged from care and identify detailed patterns of behavior that can help target interventions to improve retention in the first year of treatment.

### Methods

#### Study populations and data

Sites for this study were public sector, primary healthcare clinics in Gauteng, Mpumalanga, and KwaZulu Natal (KZN) provinces in South Africa. Data were drawn from six facilities in West Rand District in Gauteng, six in Ekhureleni District in Gauteng, six in Enhlanzeni District in Mpumalanga, and six in King Cetshwayo District in KZN. The six sites in each district were selected because they were relatively large and captured diversity in facility setting (urban v rural) and in nongovernmental support partners. Access to data required both facility- and district-level department of health approvals, along with institutional ethics review and National Department of Health authorization.

For this study, we analyzed anonymized, retrospective cohort data from South Africa’s national electronic medical record system, Tier.net[13]. Individual, longitudinal HIV treatment records included site, date of ART initiation, observed dates of clinic visits and next scheduled clinic visit, sex, age at initiation, and observed dates of death or transfer to another facility. Scheduled visit dates reflected the healthcare provider’s determination of when a client should next interact with the healthcare system for a clinical consultation and/or medication refill.

Inclusion criteria for the analytic sample were 1) observed ART initiation visit on or after 1 January 2018; and 2) ≥ 14 months of potential follow-up reflected in the dataset prior to the censor date for outcome variables. The 14-month endpoint provided a two-month window to observe visit attendance occurring after our 12-month observation period so that we could correctly classify visits scheduled at 12 months. Records from clients aged <18 years at date of ART initiation and records with a recorded ART initiation date but no observed visit dates in the datasets—including no visit on reported date of ART initiation—were excluded. Dataset censoring dates were 9 January 2023 for Mpumalanga, 6 April 2022 for KwaZulu Natal, and 8 April 2021 for Gauteng sites and represent the date of the latest visit observed in each of the datasets. Differences in censor dates reflect when the data were provided to the study team and which records and fields were made available to us in each district.

#### Variable definitions and outcomes

For each individual in the dataset, we first categorized each observed clinic visit after the ART initiation visit into one of three categories in relation to the next scheduled clinic visit date entered into the medical record before the visit in question: 1) “as planned” for visits that occurred on or before the next scheduled date; 2) “attended late ≤28 days” for visits that were attended after but within 28 days of the scheduled date; and 3) “attended late >28 days” for visits that were attended more than 28 days after the scheduled date (Table 1). We also defined a “scheduled visit not attended” on the date at which disengagement was recognized for scheduled visits that were not ever attended. The scheduled visit not attended date was defined for any particular client as 28 days after the last scheduled visit date.

**Table 1:** Visit types defined for study.

| Visit category | Definition |
| --- | --- |
| Initiation visit | Observed date of ART initiation |
| Attended as planned | Visits attended on or before the next scheduled date |
| Attended late $\leq 28$ days | Visits attended after but within 28 days of the scheduled date |
| Attended late $\geq 28$ days | Visits attended more than 28 days after the scheduled date |
| Scheduled visit not attended | Date at which disengagement was recognized after a scheduled visit was missed.<br>This date is 28 days after the last scheduled, unattended visit. |

We then used these visit classifications to specify a set of engagement patterns (outcomes) for the periods from 0-6 months after initiation and from 7-12 months after initiation, as defined in Table 2 and summarized graphically in Supplementary Figure 1. Client engagement was classified at these time points into one of three categories: continuously engaged in care, cyclically engaged in care, or disengaged from care. Disengagement was further categorized by the time at which disengagement was recognized, either immediately after initiation, within the first 3 months on ART, or between month 3 and 6 on ART. Participants who attended visits throughout the observation period and had a next scheduled visit date after 365 days were considered to be engaged in care at 12 months, even if no visit was observed in the data set after the 365-day mark.

**Table 2.** Patterns of engagement defined for study.

| Label | Pattern of engagement observed during months 0-6 after ART initiation | Pattern of engagement observed during months 7-12 after ART initiation |
| --- | --- | --- |
| Continuous | No visit > 28 days late in first 6 months and next visit scheduled >6 months after initiation (i.e. all visits as planned or late $\leq 28$ days) | Continuous or cyclical at 6 months AND no visit > 28 days late in months 7-12 and next visit scheduled >12 months after initiation (i.e., all visits attended as planned or late $\leq 28$ days) |
| Cyclical | Attended at least 1 visit late by more than 28 days between initiation and month 6 but subsequently re-engaged in care by 6 months after initiation. | Continuous or cyclical at 6 months; attended at least one visit late by more than 28 days between months 7 and 12 but subsequently re-engaged in care by 12 months after initiation. |
| Immediate disengagement | No visits after date of ART initiation; not observed during months 0-6 after ART initiation | Not reported for months 7-12; aggregated under "Disengaged months 0-6" |
| Early disengagement | $\geq 1$ visit after date of ART initiation but last visit $\leq 3$ months after initiation | Not reported for months 7-12; aggregated under "Disengaged months 0-6" |
| Late disengagement | $\geq 1$ visit after date of ART initiation but last observed visit occurs 4-6 months after initiation (i.e. no further scheduled or observed visits in 7-12 month period) | Not reported for months 7-12; aggregated under "Disengaged months 0-6" |
| Disengaged months 0-6 | Composite pattern including all immediate, early, or late disengagers in first 6 months; no visits observed after 6 months | Classified as immediate, early, or late disengagers in first 6 months AND no visits observed during months 7-12 |
| Disengaged months 7-12 | Not reported for months 0-6 | Classified as continuous or cyclical at 6 months; at least 1 visit observed in months 7-12 but $\geq 1$ scheduled visit late by >28 days with no evidence of return during months 7-14 |
| Transferred | Documented transfer to another healthcare facility during month 0-6 | Documented transfer to another healthcare facility at any time from 0-12 months |
| Died | Death recorded at any time from 0-6 months | Death recorded at any time from 0-12 months |

#### Statistical analysis

We first report cohort and participant characteristics at ART initiation using frequencies and simple proportions stratified by province. Next, we describe the frequency and distribution of each defined visit type during the first 6 and 12 months after ART initiation as a proportion of the total monthly visits for that visit type, stratified by sex, age, district, and year of ART initiation.

We then report the frequency and proportion of each of the engagement patterns defined at 6- and 12-months after ART initiation and characterize each pattern according to median number of visits and visit types during months 0-6 and 7-12 on ART. We illustrate these outcomes using an alluvial chart for depicting both the first and second six-month periods after ART initiation and illustrating how participants may have shifted from one engagement pattern to another between the two time periods.

We investigate potential predictors of disengagement at both 6- and 12-months on ART using crude risk ratios and their corresponding 95% confidence intervals (CI). We also adjusted each estimate for other predictors using log-linear regression models to estimate adjusted risk ratios (aRR). We then estimate the effect of engagement pattern during the first 6 months on ART on the risk of disengagement during the second 6 months on ART. As participants who had disengaged, transferred, or died during the first 6 months on ART could not be “newly” defined as disengaged during months 7-12, this analysis was restricted to the participants with engagement patterns characterized as continuous or cyclical at 6 months after staring ART. We conducted a crude analysis comparing the proportion of participants cyclically engaged at 6 months who disengaged from care by 12 months after ART initiation to the proportion of participants continuously engaged at 6 months achieving the same outcome. We estimated crude risk ratios and corresponding 95% confidence intervals and used log-linear regression models to adjust estimates for potential confounding and present adjusted risk ratios and corresponding 95% CI.

#### Outcome misclassification and sensitivity analysis

The data for this analysis are limited to within-facility observations; we could not track clients from one facility to another. It is thus likely that outcome misclassification bias may occur in our estimates due to “silent” transfers. In these instances, participants who did not attend a scheduled visit have not disengaged from care but are instead attending visits at another facility. Prior estimates indicate that outcome misclassification of this type may range from 8% [14] to 26% [15] within the context of ART treatment programs in the region. To account for the effect of outcome misclassification on our estimates, we performed four sensitivity analyses according to the different directions in which misclassification could occur: 1) we assumed equal rates of 6-month outcome misclassification for both the continuous and cyclically engaged participants at the lowest end of the range (8%); 2) we assumed equal rates of 6-month outcome misclassification for both the continuous and cyclically engaged participants at the highest end of the range (26%); 3) we assumed 6-month outcome misclassification occurred at the lower rate of 8% among continuously engaged participants and at the higher rate of 26% among cyclically engaged participants; and, finally 4) we assumed 6-month outcome misclassification occurred at the higher rate of 26% among continuously engaged participants and at the lower rate of 8% among cyclically engaged participants. Crude estimates of the effect of engagement pattern at 6 months on risk of disengagement from care by 12 months on ART accounting for potential outcome misclassification are presented as relative risks with 95% confidence intervals for each potential direction of outcome misclassification.

#### Ethics statement

Analysis of the de-identified datasets was approved by the University of the Witwatersrand Human Research Ethics Committee (Medical) under protocol M1902105 and by the Boston University Medical Center IRB under protocol H-38815 for the use of data with a waiver of consent. All data were de-identified prior to sharing with the study teams and no direct participant contact occurred.

## Results

### Description of study population

Our analytic sample included a total of 33,821 ART clients. Study site, demographic, and HIV treatment characteristics are shown in Table 3, by province and district. Cohorts were similar in terms of proportion female (between 64%-68%) and median age at ART initiation (between 31-33 years).

**Table 3.** Characteristics of study cohorts stratified by district (n=33,821)*

| Characteristic | Mpumalanga cohort | KZN cohort | Gauteng cohorts |  |
| --- | --- | --- | --- | --- |
| District | Enhlanzeni | King Cetshwayo | West Rand | Ekurhuleni |
| Total sample (n) | 21,119 | 11,281 | 5,961 | 13,188 |
| Dataset censoring date | 09 Jan 2023 | 06 Apr 2022 | 09 Jan 2021 | 8 Apr 2021 |
| Included in analytic sample (n, %)* | 12,813 (61%) | 8,741 (77%) | 3,695 (62%) | 8,572 (65%) |
| Number of study sites | 6 | 6 | 6 | 6 |
| Female (n, %) | 8,136 (63.5%) | 5,959 (68.2%) | 2521 (68.2%) | 5651 (65.9%) |
| Age at ART initiation |  |  |  |  |
| Median (IQR) | 32 [27, 40] | 31 [25, 37] | 33.0 [27, 41] | 33.0 [28, 40] |
| 18-25 | 2,595 (20.3%) | 2,209 (25.3%) | 661 (17.9%) | 1,413 (16.5%) |
| 26-49 | 9,150 (71.4%) | 6,012 (68.8%) | 2,703 (73.2%) | 6,520 (76.1%) |
| 50+ | 1,068 (8.3%) | 520 (5.9%) | 331 (9.0%) | 639 (7.5%) |
| Year of ART initiation |  |  |  |  |
| 2018 | 2,097 (16.4%) | 3,187 (36.5%) | 2,106 (57.0%) | 3,700 (43.2%) |
| 2019 | 3,819 (29.8%) | 3,273 (37.4%) | 1,589 (43.0%) | 4,322 (50.4%) |
| 2020 | 4,154 (32.4%) | 2,090 (23.9%) | 0 (0%) | 550 (6.4%) |
| 2021 | 2,743 (21.4%) | 191 (2.2%) | 0 (0%) | 0 (0%) |
| Number of facility visits between initiation and 12 months on ART (median, IQR)† | 6 (2, 9) | 9 (5, 12) | 8 (4, 10) | 7 (3, 10) |
| Time between initiation and last visit or defined date of disengagement (days) (median, IQR) | 372 (113, 408) | 390 (264, 411) | 383 (199, 411) | 374 (140, 402) |

\*Analytic sample was restricted to client records with ART initiation dates on or after 1 January 2018 and a minimum of 14 months' potential observation time.
†Inclusive of the visit at which ART was initiated

The median number of visits observed during the first 12 months on ART, including the visit at which ART was initiated, was highest in the KZN sites (9 visits) and lowest in the Mpumalanga sites (6 visits); interquartile ranges were broad, from a low of just 2 visits to a high of 12.

### Distribution of visit types in first 6 and 12 months on ART

A total of 228,765 clinic visits were observed during the first 12 months on ART and classified into the five visit types shown in Table 1 (Supplementary table 1). Most (63%) of the 115,143 visits that were attended after initiation in the first 6 months of care happened as planned, while a quarter were attended late but within 28 days. Only 4% of scheduled visits during this period were attended late by more than 28 days after the date scheduled, but 8% of scheduled visits were never attended at all. Similar results were observed when considering all visits during the first 12 months on ART.

As indicated in Table 4, the majority (63%) of 209,337 visits scheduled during the 12-month period after initiation were also attended as planned. A quarter (25%) of all first-year visits after initiation were late, but by less than 28 days. Only 6% were late by more than 28 days, while 7% were not attended at all. The proportion of scheduled visits not attended peaked sharply in month 2 after ART initiation at 18%, some three times higher than in any other month during the first year on ART (Supplementary figure 2).

**Table 4.** Classification of visit types during the first six and twelve months after ART initiation, by engagement pattern.

|  | Engagement pattern months 0-6 after ART initiation |  |  |  |  |  |  | Total |
| --- | --- | --- | --- | --- | --- | --- | --- | --- |
|  | *Continuous* | *Cyclical* | *Immediate*<br>*disengagement* | *Early*<br>*disengagement* | *Late*<br>*disengagement* | *Transferred* | *Died* |  |
| N (clients) | 19,382<br>(57.3%) | 4,835<br>(14.3%) | 3,106<br>(9.2%) | 531<br>(1.6%) | 1,721<br>(5.7%) | 3,841<br>(11.4%) | 405<br>(1.2%) | 33,821<br>(100%) |
| Number of visits per client in first 6 months after initiation (median, IQR) | 5 [2, 6] | 4 [3, 5] | 1 [1, 1] | 2 [2, 2] | 3 [2, 4] | 2 [1, 3] | 2 [1, 3] | 4 [2, 6] |
| Total visits in first six months after initiation* | 83,813 | 12,922 | 3,106 | 1,093 | 5,093 | 8,246 | 870 | 115,143 |
| Attended as planned | 72.7% | 44.3% | 0% | 32.4% | 39.6% | 34.1% | 36.1% | 62.7% |
| Attended late <28 days | 27.3% | 22.3% | 0% | 18.7% | 21.3% | 15.8% | 15.9% | 24.7% |
| Attended late >28 days | 0% | 33.4% | 0% | 0.4% | 5.3% | 3.5% | 1.5% | 4.2% |
| Scheduled visit in first 6 months not attended# | 0% | 0% | 100% | 48.6% | 33.8% | 46.6% | 46.6% | 8.3% |
|  | Engagement pattern months 7-12 after ART initiation |  |  |  |  |  |  | Total |
|  | *Continuous* | *Cyclical* | *Disengaged 0-*<br>*6 months* | *Disengaged 7-*<br>*12 months* | *n.a.* | *Transferred* | *Died* |  |
| N (clients) | 14,941<br>(44.2%) | 5,110<br>(15.1%) | 5,358 (15.8%) | 2,450 (7.2%) |  | 5,443<br>(16.1%) | 523 (1.5%) | 33,821<br>(100%) |
| Number of visits attended per client in first 12 months after initiation (median, IQR) | 10 [8, 11] | 8 [6, 10] | 1 [1, 2] | 5 [4, 6] |  | 3 [1, 5] | 2 [1, 4] | 7 [3, 10] |
| Total visits in first 12 months after initiation* | 131,847 | 35,644 | 9,292 | 12,639 |  | 18,300 | 1,615 | 209,337 |
| Attended as planned | 71.9% | 51.6% | 25.5% | 45.8% |  | 43.6% | 44.9% | 62.9% |
| Attended late <28 days | 26.6% | 28.9% | 13.9% | 25.6% |  | 21.4% | 18.6% | 25.0% |
| Attended late >28 days | 1.5% | 19.5% | 2.9% | 9.3% |  | 5.8% | 4.2% | 5.5% |
| Scheduled visit not attended# | 0% | 0% | 57.7% | 19.4% |  | 29.2% | 32.3% | 6.6% |

\*See Table 1 for definition of visit types
#Not included for calculation of number of visits attended in row above.

### Engagement patterns 6 and 12 months after ART initiation

At the 6-month endpoint, 57% of the cohort remained continuously engaged, 14% showed a pattern of cyclical engagement, and 16% had disengaged from care (Figure 1 and Table 4).

**Figure 1.**
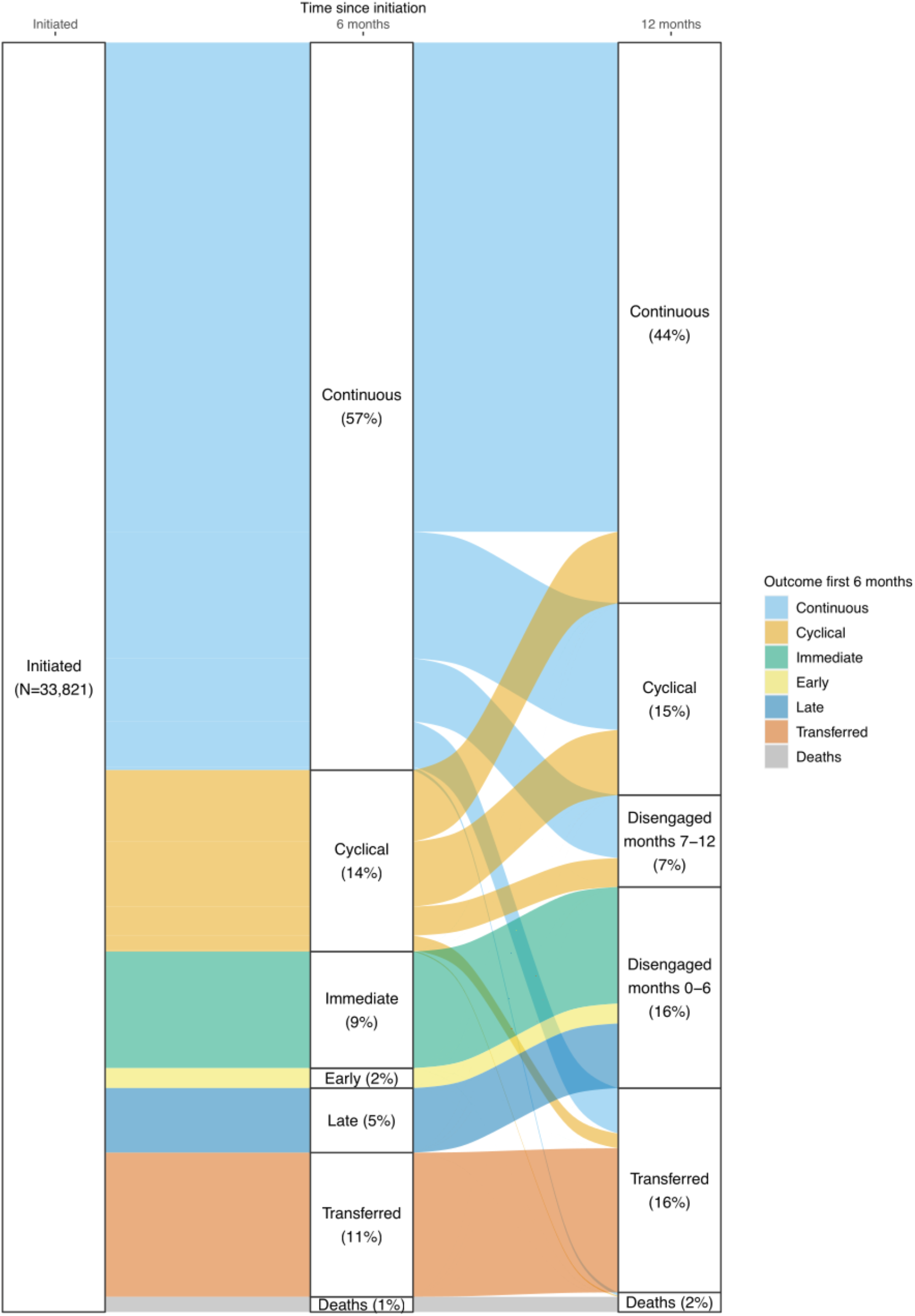
Engagement patterns at 6 and 12 months after ART initiation.

Among the 16% who disengaged from care, 58% disengaged immediately after ART initiation, 10% within the first 3 months on ART (early disengagers), and 32% between months 3 and 6 after initiation (late disengagers). Transfers occurred among 11% of the cohort, and the remaining 1% were reported to have died by 6 months after initiation. Participants who had continuously engaged in care at 6 months attended nearly three quarters (73%) of their visits as planned.

Those with cyclical engagement attended fewer than half of their scheduled clinic visits as planned (44%), while those who disengaged from care during the first 6 months on ART attended only a third of visits as planned (32% among early disengagers and 40% among late disengagers).

Table 5 reports combined engagement patterns observed at 6- and 12-months after ART initiation, results also illustrated in Figure 1. At 12 months after initiation, most of those who were continuously engaged at 6 months (67.2%), stayed in that category, but a substantial minority (17.4%) shifted to a cyclical engagement pattern, while others (8.7%) disengaged from care during this period. Most who were cyclically engaged during the first six months shifted to continuous care (39%), with some remaining cyclical in their engagement (35.9%) and some becoming disengaged (15.9%). At the 12-month endpoint, a total of 44.4 % had been continuously engaged throughout the period from months 7 to 12, 15.1% ended as cyclical engagers, 16.1% had transferred to another facility, 1.5% had died, and 7.2% fell into the 7-12 month disengagement category.

**Table 5.**
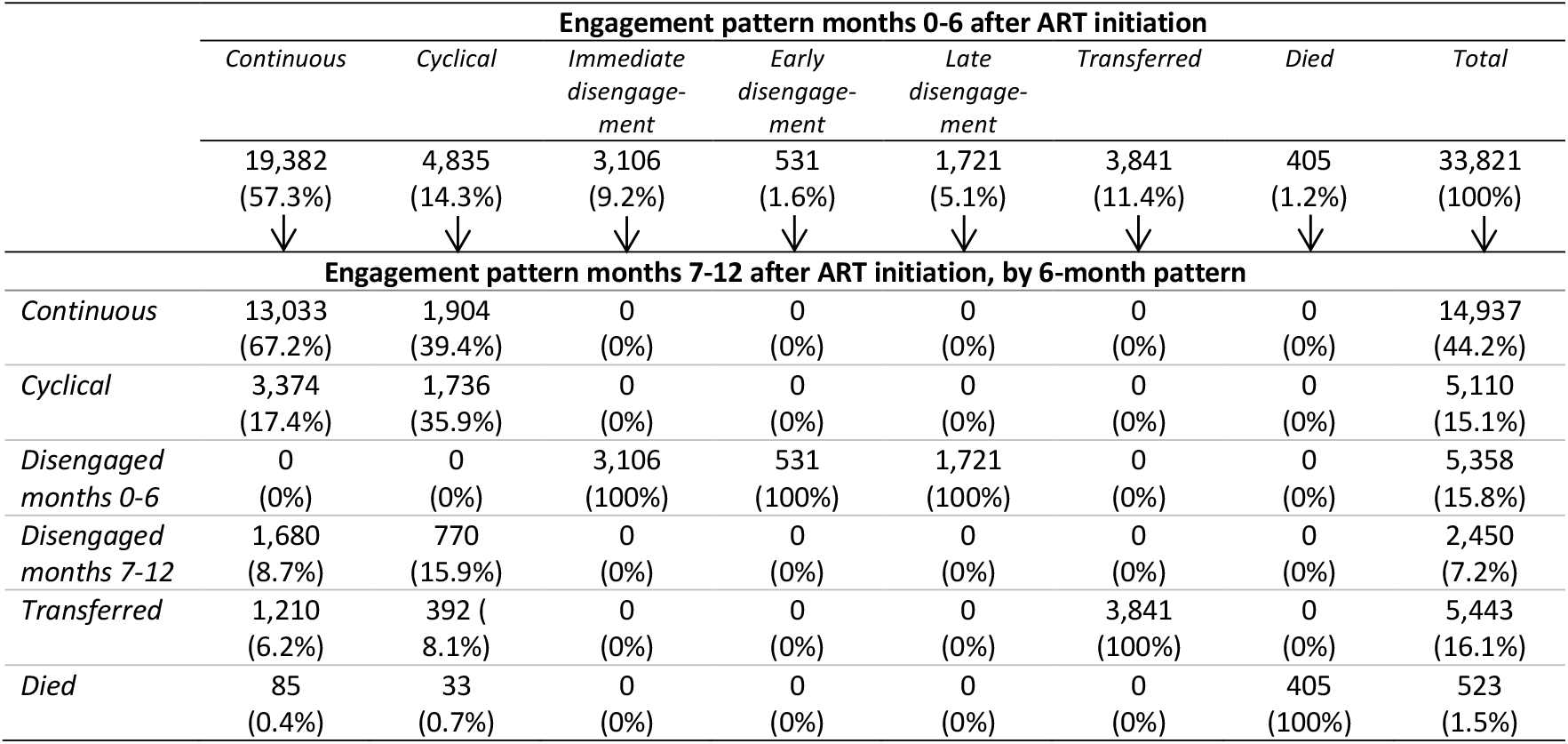
Engagement patterns 6 and 12 months after ART initiation.

|  | Engagement pattern months 0-6 after ART initiation |  |  |  |  |  |  | Total |
| --- | --- | --- | --- | --- | --- | --- | --- | --- |
|  | *Continuous* | *Cyclical* | *Immediate disengagement* | *Early disengagement* | *Late disengagement* | *Transferred* | *Died* |  |
|  | 19,382<br>(57.3%) | 4,835<br>(14.3%) | 3,106<br>(9.2%) | 531<br>(1.6%) | 1,721<br>(5.1%) | 3,841<br>(11.4%) | 405<br>(1.2%) | 33,821<br>(100%) |
|  | ↓ | ↓ | ↓ | ↓ | ↓ | ↓ | ↓ | ↓ |
| Engagement pattern months 7-12 after ART initiation, by 6-month pattern |  |  |  |  |  |  |  |  |
| *Continuous* | 13,033<br>(67.2%) | 1,904<br>(39.4%) | 0<br>(0%) | 0<br>(0%) | 0<br>(0%) | 0<br>(0%) | 0<br>(0%) | 14,937<br>(44.2%) |
| *Cyclical* | 3,374<br>(17.4%) | 1,736<br>(35.9%) | 0<br>(0%) | 0<br>(0%) | 0<br>(0%) | 0<br>(0%) | 0<br>(0%) | 5,110<br>(15.1%) |
| *Disengaged months 0-6* | 0<br>(0%) | 0<br>(0%) | 3,106<br>(100%) | 531<br>(100%) | 1,721<br>(100%) | 0<br>(0%) | 0<br>(0%) | 5,358<br>(15.8%) |
| *Disengaged months 7-12* | 1,680<br>(8.7%) | 770<br>(15.9%) | 0<br>(0%) | 0<br>(0%) | 0<br>(0%) | 0<br>(0%) | 0<br>(0%) | 2,450<br>(7.2%) |
| *Transferred* | 1,210<br>(6.2%) | 392<br>(8.1%) | 0<br>(0%) | 0<br>(0%) | 0<br>(0%) | 3,841<br>(100%) | 0<br>(0%) | 5,443<br>(16.1%) |
| *Died* | 85<br>(0.4%) | 33<br>(0.7%) | 0<br>(0%) | 0<br>(0%) | 0<br>(0%) | 0<br>(0%) | 405<br>(100%) | 523<br>(1.5%) |

By one year after initiation, only 38.5% of the cohort had maintained continuous engagement for the full 12 month period. An additional 5.1% had engaged cyclically for the entire year. 15.6% shifted between continuous and cyclical engagement over the course of the year. Overall disengagement during the first year totaled 23%.

### Patterns of engagement by province, year of initiation, age, and gender

Patterns of engagement in the first 6 months of care differed slightly between provinces. KwaZulu-Natal had the lowest proportion of client disengagement at 10%, compared to 17% in Mpumalanga and 19% and 15% in the two Gauteng districts (Supplementary table 2). Differences by province followed similar trends for months 7-12 on ART as in the first 6 months.

Disengagement between months 7 to 12 was highest in Gauteng (EK: 10%; WR: 11%), followed by Mpumalanga (6%) and KwaZulu-Natal (5%). Cyclical engagement increased in Mpumalanga (10%) and KwaZulu-Natal (20%) but decreased in Gauteng (17%). The number of visits per participant was highest in KwaZulu-Natal (9, IQR 5-12), followed by Gauteng (EK: 7, ICR 3-10; WR: 8, ICR 4-10) and Mpumalanga (6, IQR 2-9), respectively.

Annual trends in the patterns of engagement also differed between provinces. In Mpumalanga, disengagement at 6 months increased from 12% among participants initiated in 2019 to 18% in 2020 and 17% in 2021, respectively. Rates of cyclical engagement in Mpumalanga trebled between 2019 (4%) and 2021 (14%). In KwaZulu Natal, however, continuous engagement in care increased from 52% in 2018 to 64% in 2019 and 70-71% in 2020-2021. In the Ekurhuleni district of Gauteng, continuous engagement decreased by nearly 10% over the study period (2018-2020), but due to increases in the proportion of participants who transferred to other facilities, disengagement decreased by 2% overall, meaning overall engagement improved. Deaths remained stable or were lower at both end points in 2020 compared to earlier and later years, despite the COVID-19 pandemic. In general, participants who initiated ART treatment in 2020 had the lowest rates of disengagement (20%) at 12 months and showed improvement from 2018 (27%) and 2019 (21%).

Age was strongly associated with differences in engagement profile (Table 6). The proportion of participants engaged in a continuous pattern of care during the first 6 months rose from 48% among participants aged 18-25 to 59% among those aged 25-49 years to 66% for those aged ≥50 years. Participants aged 18-25 years were most likely to be engaged in a cyclical pattern (17% compared to 14% (26-49) and 11% (≥50 years)) or have transferred to a different facility (15% compared to 11% and 9%, respectively). Increased age was associated with a higher proportion of participants dying in the first year after ART initiation (0.4% among 18-25 vs 3% for those aged ≥50 years). Patterns of engagement in months 7-12 also indicated differences by age group. Younger participants (aged 18-25 years) were less likely to be engaged in continuous care than those aged 25-49 years and those ≥50 years (34% vs 46% and 55% respectively). Those aged 18-25 years were also more likely to be engaged in cyclical care (16% vs 15% and 12%), to have disengaged from care (28% vs 23% and 16%), or to have transferred (22% vs 15% and 11%) than those age 25-49 years and those ≥50 years, respectively. Participants aged 18-25 also attended fewer visits (5; IQR 1-8) than did their older counterparts (26-49 years: 6 visits (IQR 2-9); ≥50 years: 7 visits (IQR 3-9)).

**Table 6:**
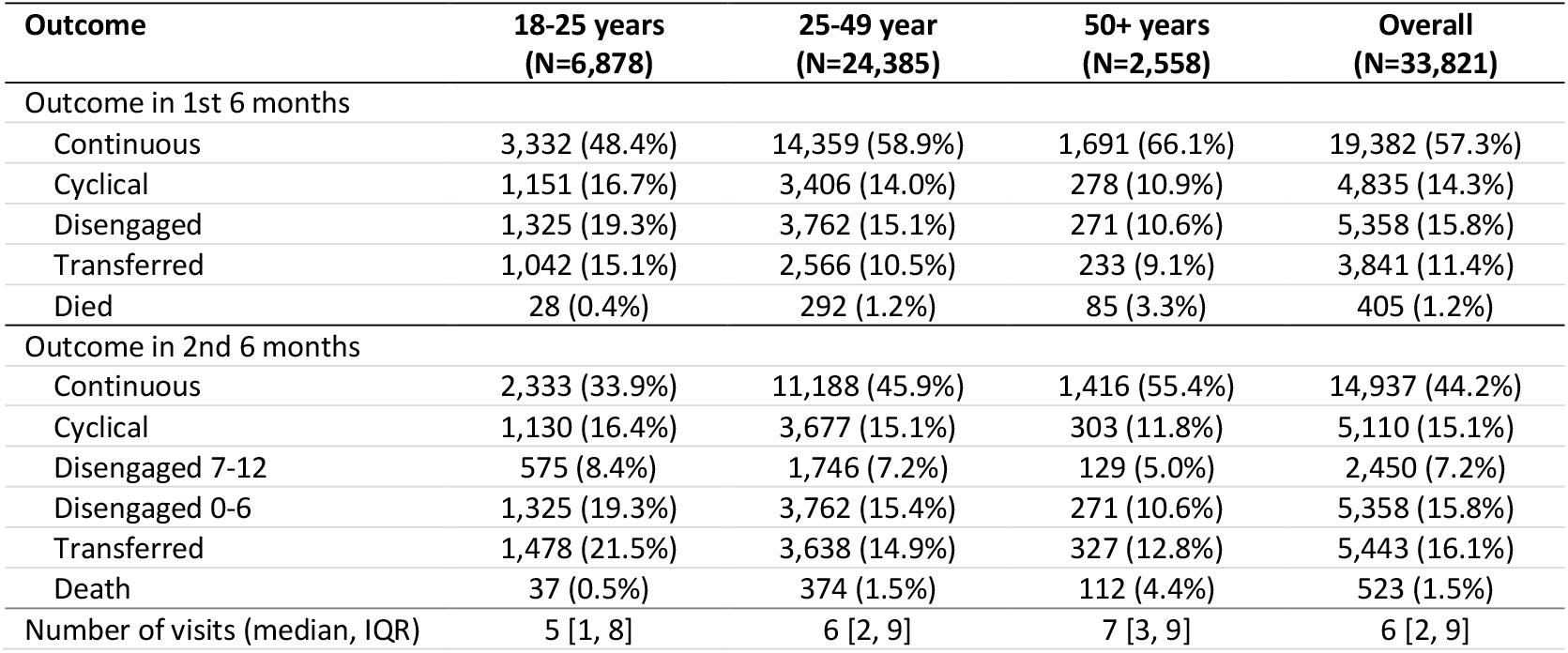
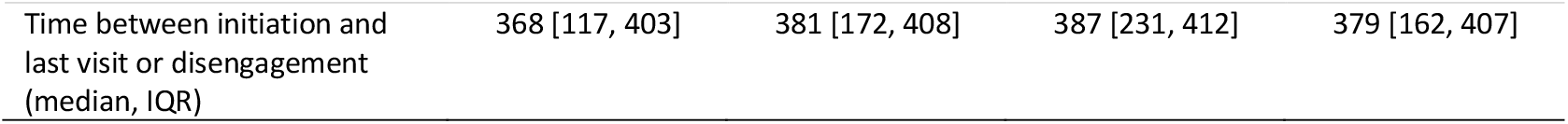
Classification of visit types and engagement patterns during the first and second six months after ART initiation stratified by age group.

No meaningful differences in engagement pattern were observed between females and males during either the first or second 6-month periods on ART (Supplementary table 3).

### Predictors of outcomes and sensitivity analysis

As noted above, age was strongly associated with risk of disengagement from in the first and second 6-month periods on ART (Supplementary table 4). Participants aged 18-25 years were 1.82 (95% CI: 1.61-2.06) times more likely to disengage than those older than 50 years, while those aged 26-49 were 1.46 (1.30-1.64) times more likely to be disengaged at month 6. Differences in risk of disengagement by age remained after adjusting for gender, year of ART initiation, and district, with those aged 18-25 being nearly twice (RR 1.97, 95% CI: 1.74-2.23) as likely to be disengaged and those aged 26-40 nearly at 50% (RR 1.49, 95% CI: 1.32-1.67) higher risk of being disengaged at 6 months when compared to those older than 50 years. Participants who initiated in 2019 and 2020 had lower risk of disengagement in the first 6 months (crude RR: 0.72) compared to 2018. This finding persisted when adjusting for age, district, and gender (2019 adjusted RR: 0.71, 95% CI: 0.67-0.78; 2020: RR 0.73(95% CI: 0.67-0.78)).

When considering disengagement between months 7-12, those who were engaged cyclically in months 0-6 had 1.8 (95% CI: 1.70-1.99) times the risk of becoming disengaged in months 7-12 compared to those who were continuously engaged in months 0-6. This pattern persisted when adjusting for age, gender, year of initiation, and district (adjusted RR 1.71; 95% CI: 1.58-1.86).

Age was also associated with higher risk of disengagement: those aged 18-25 years were about twice(crude RR: 1.95 (95% CI: 1.63-2.23) adjusted RR: 2.09 (95% CI: 1.74-2.51) as likely as those older than 50 years to become disengaged.

In sensitivity analysis, we observed the impact of potential outcome misclassification on the estimates of risk of disengagement due to silent transfer (Supplementary table 5). If we were to assume non-differential misclassification (misclassification of outcomes happened in equal proportions to both the continuous engagers and cyclical engagers), our original analysis would underestimate risk of disengagement by 10 percentage points regardless of the extent of misclassification. Under the assumption of differential misclassification (i.e. misclassification was more likely to happen to either the group classified as continuous engagers or the group classified as cyclical engagers), the impact of misclassification was greater and results differed depending on the direction of misclassification. If misclassification were to be more frequent among cyclical engagers, our original analysis would underestimate risk of disengagement by nearly 50 percentage points. If misclassification were to be more frequent among continuous engagers, our original analysis would overestimate risk of disengagement by nearly 40 percentage points.

## Discussion

In this study of more than 30,000 clients initiating HIV treatment in four districts in South Africa, fewer than 60% and 45% of clients were continuously engaged in care (no interruptions >28 days) at 6 and 12 months after treatment initiation, respectively, at their initiating facilities.

Cyclical engagement, a pattern of remaining in care but being late for visits, suggesting interruptions in medication adherence, was common, experienced by some 14% of clients even in their first six months. Nearly 90% of participants who disengaged from care during the first 6 months on ART did so either immediately after the initiation visit or within the first three months on ART. Almost a quarter of clients in the cohort (23%) had disengaged from ART by the end of the first year.

Continuity of treatment remains a critical objective in ending the HIV epidemic. Despite huge advances in access to ART in the era of UTT, current models of HIV service delivery still struggle to achieve high levels of sustained engagement in care after treatment initiation, particularly in the early treatment period. Our results suggest that not only is the first 6 months on ART a high-risk period for disengagement from care; it also appears to be critical for establishing long terms patterns of engagement with HIV care programs. For the most part, patterns of engagement that were established during the first 6 months on ART demonstrated little change in months 7-12.

Cyclical and disengagement pattern outcomes by 6 months on ART, moreover, were frequently preceded by multiple occurrences of visits attended late, suggesting opportunities to intervene with clients at risk of disengagement may be missed in the early ART period if visit patterns are not recognized. Given how many of the clients (58%) who disengage from care in the first six months do so immediately after ART initiation, waiting for a second visit to intervene is too late.

The six-month outcomes we observed are consistent with those reported by others in South Africa and in the region more broadly. A systematic review of retention on HIV treatment estimated an average 6-month retention rate of 85% for studies from in South Africa[16], while a more recent analysis of routinely collected data reported that 74% of participants in South Africa were still engaged at 6 months after initiation[2]. Our study adds to the existing literature by examining the early treatment period in a level of detail not previously reported and relating patterns of engagement to visit behavior. One exception to the consistency of engagement patterns over the first year on ART was observed among those with a pattern of cyclical engagement in care. Despite not fully disengaging from care, participants with cyclical patterns of care were less likely to attend scheduled visits as planned compared not only to those engaging in care continuously but to those who fully disengaged from care within the first 6 months on treatment. As mentioned above, irregular visit attendance likely means at least some periods of treatment interruption during this critical period on ART, potentially compromising the client’s own health[17] and creating opportunities for ongoing transmission of HIV that can be prevented with sustained ART [18]. While interruptions are increasingly being seen as “normal” for lifelong chronic disease treatment[19], they are still not desirable, particularly before an ART client has achieved sustained viral suppression[17].

Cyclical engagement as we observe it in these datasets may reflect new initiators’ need for greater flexibility in access to care, as has been demonstrated by differentiated service delivery (DSD) models throughout the region[20–22]. In South Africa, as in many other countries, however, eligibility for access to differentiated models of care has historically been limited those already on ART for at least 6 months [23]. Those in the first 6 months continue to be required to make multiple clinic visits and given just 1 or 2 months’ supplies of medications at a time. The sites in this study, for example, informed study staff that during the study period they asked clients to come to the clinic an average of 7 or 8 times in the first 6 months, including the visit at which treatment is initiated. We have found similar practices in Zambia, another high HIV-prevalence southern African country facing challenges with retention in care[24].

In our study, nearly 30% of individuals who demonstrated cyclical patterns of engagement during the first 6 months on ART were observed to engage continuously during months 7-12, a period when they were eligible for less burdensome DSD models such as community-based medication pickups. In recognition of the potential value of earlier eligibility for DSD models, in April 2023 the South African National Department of Health announced that going forward, viral load tests will be performed after 3 months on ART rather than 6, and clients with viral suppression will be eligible for enrollment in DSD models at the 4-month point[25]. While this will not immediately affect the very early disengagers seen in our study, it may encourage some to persist in care for the short period until they are eligible.

We observed the lowest rates of continuous engagement in care among youth aged 18-25 years, with less than half continuously engaged in care at 6 months and only a third continuously engaged by 12 months. Concerning rates of disengagement form care on ART among adolescents and youth have been described previously [26–30]. Despite implementation of models of care designed to address retention challenges in this group, such as school and youth adherence clubs, youth retention remains sub-optimal. Understanding reasons for disengagement among young adults will be critical if we are to design effective interventions that can affect patterns of engagement from the start of HIV treatment [31].

Our results should be considered within the context of the limitations relevant to observational cohorts comprised from routinely collected EMR sources. First, the datasets analyzed here do not observe clinic attendance outside of the facilities at which participants initiated ART and are thus not robust to silent transfers. Rather than speculate on the possible impact of this type of outcome misclassification, we performed sensitivity analyses considering scenarios of both differential and non-differential misclassification of disengagement from care. The results suggest that non-differential misclassification (more likely in this context as both exposure and outcome variables were assessed independently of each other) would result in underestimation of the risk of disengagement from care: even in the presence of poor sensitivity of outcome misclassification, the estimate of the effect of pattern of engagement at 6 months on ART on subsequent disengagement in care was likely biased only minimally towards the null.

Second, as we noted previously, visit attendance is used here as a proxy for medication adherence but may not directly correlate. To the extent to which visit attendance was more consistent than medication adherence, our estimates would over-estimate engagement in care. On the other hand, the EMR datasets we used may not observe medication pickups at external pickup points, which may have been utilized by some participants during the second 6 months on ART. While using scheduled visit dates in relation to observed visits should mitigate this limitation, we cannot be sure that this occurred consistently and universally across all sites and datasets. Misclassification like this could result in over-estimation of disengagement during months 7-12. Third, dataset censoring dates differed by district, as noted above, and could have influenced results, particularly in the case of the West Rand District, where data were limited to ART initiations prior to 2020 and thus do not reflect COVID-19 pandemic impacts. Finally, restrictions on transport, clinic closures, and other COVID-19 pandemic measures likely limited access to clinic visits in the public sector starting in March 2021, such that observed engagement patterns may not reflect non-pandemic trends.

Despite these limitations, our results highlight important considerations for models of HIV service delivery during the early treatment period. We demonstrate clearly distinct patterns of engagement in HIV care during the early treatment period and their associated risk of subsequent treatment interruption. It is likely that the needs of continuous and cyclical engagers differ from those who disengage from care during the first 6 months after initiation and require different interventions or models of care. Qualitative research with persons with both continuous and cyclical patterns of engagement may reveal potentially modifiable barriers to improving outcomes in the early treatment period and beyond.

## Data Availability

Data used in this study, which were abstracted from South Africa's electronic medical record system, TIER.Net, are owned by the National Department of Health and cannot be shared by the authors. Instructions for requesting data directly from the owner of the dataset can be found at https://www.tbhivinfosys.org.za/wp-content/uploads/2022/01/Acess-to-Data-Sourced-from-the-TB_HIV-Dataset-Guidance_Final_signed.pdf.

## Supplementary files

Supplementary Table 1: Distribution of visit types during the first 12 months on ART

Supplementary Table 2: Classification of engagement profile by district and year of initiation

Supplementary Table 3: Classification of visit types and engagement patterns during the first and second six months after ART initiation, stratified by gender

Supplementary Table 4: Crude and adjusted predictors of becoming disengaged from care

Supplementary Table 5: Sensitivity analysis adjusting for potential outcome misclassification

Supplementary Figure 1: Engagement patterns and outcome definitions: 1) Continuous engagement; 2) Cyclical engagement; 3) Disengagement

Supplementary Figure 2: Distribution of last treatment visits by month after initiation

## Authors’ contributions

MM conceptualized the study, analyzed the data, and contributed to writing the original draft of the manuscript. MB conceptualized the study, analyzed the data, and contributed to writing the original draft of the manuscript. AH curated the data and contributed to the analysis. SP curated the data and contributed to the analysis. LS contributed to the analysis. All authors reviewed and edit the final manuscript.

