## Supplementary files for "Patterns of retention in care during clients’ first 12 months after HIV treatment initiation in South Africa: a retrospective cohort analysis using routinely collected data"

**Supplementary Table 1: Distribution of visit types during the first 12 months on ART**

| **Month** | **As planned** | | **Late ≤28 days** | | **Late >28 days** | | **Scheduled visits not attended** | | **Total visits** |
| --- | --- | --- | --- | --- | --- | --- | --- | --- | --- |
| Initiation |  |  |  |  |  | |  |  | 33,821 |
| Month 1 | 18,738 | 67% | 7,709 | 28% | 359 | 1% | 1,082 | 4% | 27,888 |
| Month 2 | 12,079 | 56% | 4,667 | 22% | 952 | 2% | 3,961 | 18% | 21,659 |
| Month 3 | 13,692 | 65% | 5,168 | 24% | 1,042 | 3% | 1,245 | 6% | 21,147 |
| Month 4 | 10,963 | 62% | 4,305 | 25% | 974 | 3% | 1,308 | 7% | 17,550 |
| Month 5 | 10,313 | 63% | 3,947 | 24% | 1,003 | 3% | 1,029 | 6% | 16,292 |
| Month 6 | 12,021 | 65% | 4,610 | 25% | 1,005 | 3% | 971 | 5% | 18,607 |
| Month 7 | 8,304 | 59% | 3,887 | 28% | 924 | 4% | 943 | 7% | 14,058 |
| Month 8 | 7,073 | 60% | 3,156 | 27% | 875 | 4% | 698 | 6% | 11,802 |
| Month 9 | 6,739 | 60% | 2,979 | 27% | 826 | 4% | 654 | 6% | 11,198 |
| Month 10 | 6,228 | 59% | 2,812 | 27% | 832 | 4% | 604 | 6% | 10,476 |
| Month 11 | 5,690 | 60% | 2,597 | 27% | 747 | 5% | 503 | 5% | 9,537 |
| Month 12 | 9,334 | 63% | 3,898 | 26% | 897 | 6% | 601 | 4% | 14,730 |
| Total | 12,1174 | 62% | 49,735 | 25% | 10,436 | 6% | 13,599 | 7% | 228,765 |

**Supplementary table 2. Engagement profiles by province and year of initiation**

| Outcome | Mpumalanga | | | | | KwaZulu-Natal | | | | | | Gauteng (Ekurhuleni) | | | | | | Gauteng (West Rand) | | | | |
| --- | --- | --- | --- | --- | --- | --- | --- | --- | --- | --- | --- | --- | --- | --- | --- | --- | --- | --- | --- | --- | --- | --- |
|  | 2018 | 2019 | 2020 | 2021 | Overall | | | 2018 | 2019 | 2020 | 2021 | | Overall | 2018 | | 2019 | 2020 | | Overall | 2018 | 2019 | Overall |
| Total N | 2,097 | 3,819 | 4,154 | 2,743 | 12,813 | | | 3,187 | 3,273 | 2,090 | 191 | | 8,741 | 3,700 | | 4,322 | 550 | | 8,572 | 2,106 | 1,589 | 3,695 |
| Outcome in 1st 6 months | | | | | | |  | | | | | | | |  | | | | | | | |
| Continuous | 1,024 (48.8%) | 2,409 (63.1%) | 2,432 (58.5%) | 1,557 (56.8%) | 7,422 (57.9%) | | | 1,659 (52.1%) | 2,099 (64.1%) | 1,478 (70.7%) | 133 (69.6%) | | 5,369 (61.4%) | 1,971 (53.3%) | | 2,133 (49.4%) | 247 (44.9%) | | 4,351 (50.8%) | 1,273 (60.4%) | 967 (60.9%) | 2,240 (60.6%) |
| Cyclical | 135 (6.4%) | 155 (4.1%) | 396 (9.5%) | 375 (13.7%) | 1,061 (8.3%) | | | 688 (21.6%) | 574 (17.5%) | 271 (13.0%) | 34 (17.8%) | | 1,567 (17.9%) | 749 (20.2%) | | 808 (18.7%) | 98 (17.8%) | | 1,655 (19.3%) | 303 (14.4%) | 249 (15.7%) | 552 (14.9%) |
| Disengaged | 544 (25.9%) | 447 (11.7%) | 727 (17.5%) | 479 (17.5%) | 2,197 (17.1%) | | | 516 (16.2%) | 224 (6.8%) | 131 (6.3%) | 13 (6.8%) | | 884 (10.1%) | 655 (17.7%) | | 849 (19.6%) | 85 (15.5%) | | 1,589 (18.5%) | 413 (19.6%) | 275 (17.3%) | 688 (18.6%) |
| Transferred | 341 (16.3%) | 745 (19.5%) | 545 (13.1%) | 308 (11.2%) | 1,939 (15.1%) | | | 298 (9.4%) | 339 (10.4%) | 185 (8.9%) | 10 (5.2%) | | 832 (9.5%) | 294 (7.9%) | | 482 (11.2%) | 115 (20.9%) | | 891 (10.4%) | 96 (4.6%) | 83 (5.2%) | 179 (4.8%) |
| Death | 53 (2.5%) | 63 (1.6%) | 54 (1.3%) | 24 (0.9%) | 194 (1.5%) | | | 26 (0.8%) | 37 (1.1%) | 25 (1.2%) | 1 (0.5%) | | 89 (1.0%) | 31 (0.8%) | | 50 (1.2%) | 5 (0.9%) | | 86 (1.0%) | 21 (1.0%) | 15 (0.9%) | 36 (1.0%) |
| Outcome in months 7-12 | | | | | | | | | | | | | | | | | | | | | | |
| Continuous | 864 (41.2%) | 1,953 (51.1%) | 1797 (43.3%) | 1,213 (44.2%) | 5,827 (45.5%) | | | 1,308 (41.0%) | 1,540 (47.1%) | 1,142 (54.6%) | 99 (51.8%) | | 4,089 (46.8%) | 1,507 (40.7%) | | 1,637 (37.9%) | 171 (31.1%) | | 3,315 (38.7%) | 1,004 (47.7%) | 706 (44.4%) | 1,710 (46.3%) |
| Cyclical | 91 (4.3%) | 233 (6.1%) | 616 (14.8%) | 383 (14.0%) | 1,323 (10.3%) | | | 709 (22.2%) | 648 (19.8%) | 326 (15.6%) | 50 (26.2%) | | 1,733 (19.8%) | 666 (18.0%) | | 704 (16.3%) | 107 (19.5%) | | 1,477 (17.2%) | 306 (14.5%) | 271 (17.1%) | 577 (15.6%) |
| Disengaged   0-6 months | 544 (25.9%) | 447 (11.7%) | 724 (17.4%) | 479 (17.5%) | 2,194 (17.1%) | | | 516 (16.2%) | 223 (6.8%) | 131 (6.3%) | 13 (6.8%) | | 883 (10.1%) | 655 (17.7%) | | 849 (19.6%) | 85 (15.5%) | | 1,589 (18.5%) | 413 (19.6%) | 275 (17.3%) | 688 (18.6%) |
| Disengaged  7-12 months | 113 (5.4%) | 173 (4.5%) | 249 (6.0%) | 244 (8.9%) | 779 (6.1%) | | | 140 (4.4%) | 202 (6.2%) | 97 (4.6%) | 4 (2.1%) | | 443 (5.1%) | 380 (10.3%) | | 402 (9.3%) | 40 (7.3%) | | 822 (9.6%) | 207 (9.8%) | 199 (12.5%) | 406 (11.0%) |
| Transferred | 426 (20.3%) | 933 (24.4%) | 699 (16.8%) | 394 (14.4%) | 2,452 (19.1%) | | | 474 (14.9%) | 609 (18.6%) | 361 (17.3%) | 23 (12.0%) | | 1,467 (16.8%) | 447 (12.1%) | | 671 (15.5%) | 139 (25.3%) | | 1,257 (14.7%) | 152 (7.2%) | 115 (7.2%) | 267 (7.2%) |
| Death | 59 (2.8%) | 80 (2.1%) | 69 (1.7%) | 30 (1.1%) | 238 (1.9%) | | | 40 (1.3%) | 51 (1.6%) | 33 (1.6%) | 2 (1.0%) | | 126 (1.4%) | 45 (1.2%) | | 59 (1.4%) | 8 (1.5%) | | 112 (1.3%) | 24 (1.1%) | 23 (1.4%) | 47 (1.3%) |

**Supplementary Table 3: Classification of engagement patterns during the first and second six months after ART initiation stratified by gender**

| Outcome | Female | Male |
| --- | --- | --- |
|  | (N=22,267) | (N=11,554) |
| Outcome in 1st 6 months | | |
| Continuous | 12,571 (56.5%) | 6,811 (58.9%) |
| Cyclical | 3,278 (14.7%) | 1,557 (13.5%) |
| Disengaged | 3,548 (15.9%) | 1,810 (15.7%) |
| Transferred | 2,688 (12.1%) | 1,153 (10.0%) |
| Died | 182 (0.8%) | 223 (1.9%) |
| Outcome in 2nd 6 months | | |
| Continuous | 9,700 (43.6%) | 5,241 (45.4%) |
| Cyclical | 3,362 (15.1%) | 1,748 (15.1%) |
| Disengaged 0-6 | 3,546 (15.9%) | 1,808 (15.6%) |
| Disengaged 7-12 | 1,562 (7.0%) | 888 (7.7%) |
| Transferred | 3,857 (17.3%) | 1,586 (13.7%) |
| Died | 240 (1.1%) | 283 (2.4%) |
| Number of visits (median, IQR) | 7.00 [3.00, 10.0] | 7.00 [3.00, 10.0] |
| Days between initiation and last visit or disengagement (median, IQR) | 378 [159, 407] | 380 [167, 407] |

**Supplementary Table 4: Crude and adjusted predictors of becoming disengaged from care**

| **Characteristic** | **Measure** | **Disengaged months 0-6**  **(n=5,358)** | | **Disengaged months 7-12***  **(n=2,450)** | |
| --- | --- | --- | --- | --- | --- |
|  |  | **Crude RR**  **(95% CI)** | **Adjusted RR**  **(95% CI)** | **Crude RR**  **(95% CI)** | **Adjusted RR**†  **(95% CI)** |
| 6-month outcome | Continuous | -- | -- | ref. | ref. |
|  | Cyclical | -- | -- | 1.84 (1.70-1.99) | 1.71 (1.58-1.86) |
| Age (years) | 18-25 | 1.82 (1.61-2.06) | 1.97 (1.74-2.23) | 1.95 (1.63-2.35) | 2.09 (1.74-2.51) |
|  | 26-49 | 1.46 (1.30-1.64) | 1.49 (1.32-1.67) | 1.50 (1.26-1.78) | 1.51 (1.27-1.79) |
|  | ≥50 | ref. | ref. | ref. | ref. |
| District | Gauteng (EK) | ref. | ref. | ref. | ref. |
|  | Mpumalanga | 0.92 (0.87-0.98) | 1.00 (0.93-1.06) | 0.67 (0.61-0.74) | 0.68 (0.61-0.76) |
|  | KwaZulu-Natal | 0.55 (0.51-0.59) | 0.55 (0.51-0.59) | 0.47 (0.42-0.52) | 0.47 (0.42-0.52) |
|  | Gauteng (WR) | 1.00 (0.92-1.09) | 0.97 (0.89-1.05) | 1.06 (0.95-1.19) | 1.11 (0.99-1.24) |
| Year of ART initiation | 2018 | ref. | ref. | ref. | ref. |
|  | 2019 | 0.72 (0.68-0.76) | 0.71 (0.67-0.75) | 0.96 (0.88-1.05) | 1.04 (0.95-1.13) |
|  | 2020 | 0.72 (0.67-0.78) | 0.73 (0.67-0.78) | 0.73 (0.65-0.81) | 0.98 (0.86-1.11) |
|  | 2021 | 0.87 (0.80-0.96) | 0.78 (0.71-0.86) | 1.09 (0.96-1.25) | 1.37 (1.17-1.61) |
| Sex | Female | ref. | ref. | ref. | ref. |
|  | Male | 0.98 (0.93-1.04) | 1.05 (0.99-1.10) | 1.08 (1.00-1.16) | 1.18 (1.09 – 1.28) |

**Supplementary Table 5:** **Sensitivity analysis adjusting for potential outcome misclassification**

| **Analysis** | **Crude relative risk (95% CI)** |
| --- | --- |
| **Conventional analysis (analyze data as observed)** | **1.84 (1.70-1.99)** |
| **Data adjusted for outcome misclassification:**  1. Assume non-differential outcome misclassification (8% for both) | 1.95 (1.80 – 2.11) |
| 2. Assume non-differential outcome misclassification (26% for both) | 1.95 (1.81 – 2.09) |
| 3. Assume differential outcome misclassification (8% among continuous and  26% among cyclical engagers) | 2.43 (2.25 – 2.61) |
| 3. Assume differential outcome misclassification (26% among continuous and  8% among cyclical engagers) | 1.56 (1.44 – 1.68 |

**Supplementary Figure 1:** Engagement patterns and outcome definitions: 1) Continuous engagement; 2) Cyclical engagement; 3) Disengagement

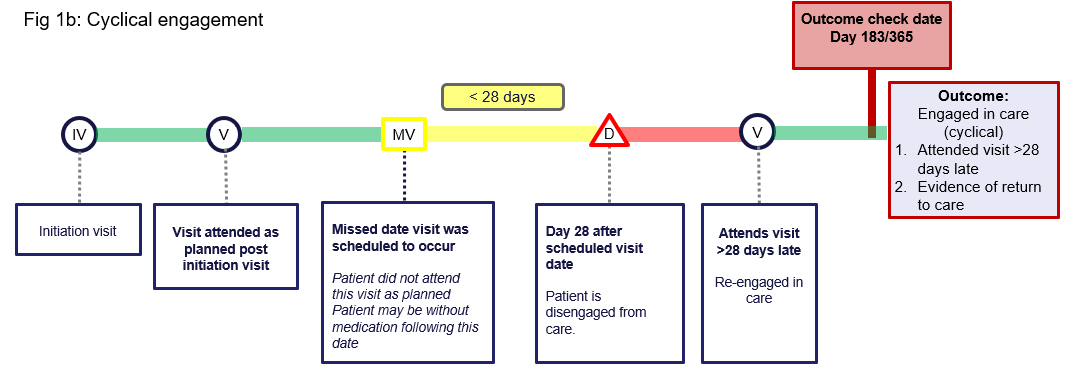

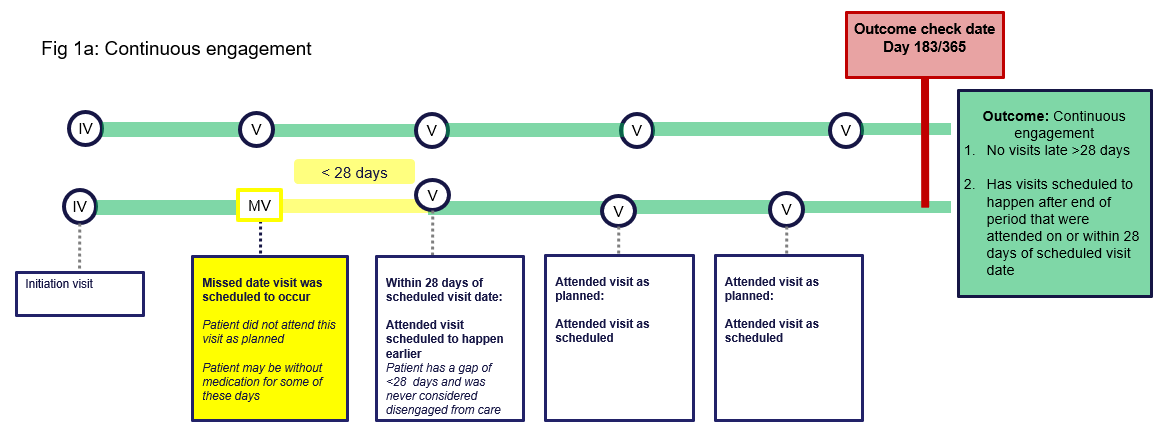

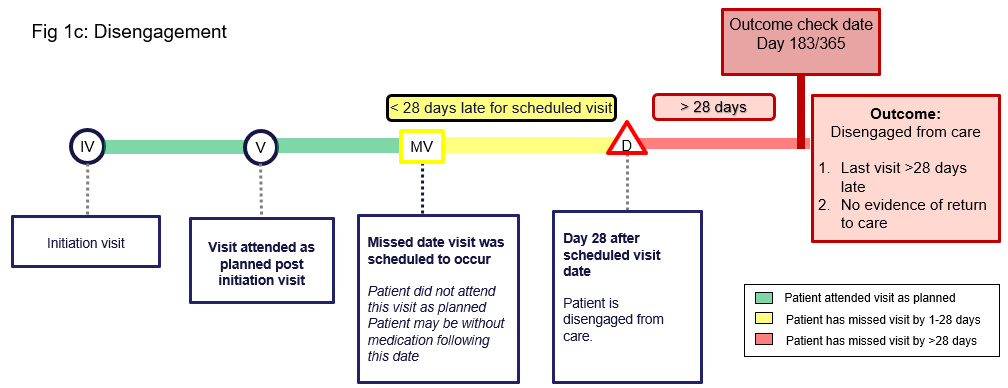

**Supplementary Figure 2**: Distribution of last treatment visits by month after initiation

|  | Visit attended late >28 days | Scheduled visits not attended |
| --- | --- | --- |
| Initiation | 0% | 0% |
| Month 1 | 1.28729% | 3.8798% |
| Month 2 | 2.33968% | 18.288% |
| Month 3 | 2.81914% | 5.88736% |
| Month 4 | 2.97696% | 7.45299% |
| Month 5 | 3.15754% | 6.31598% |
| Month 6 | 2.79717% | 5.21847% |
| Month 7 | 3.71331% | 6.70792% |
| Month 8 | 4.16139% | 5.91425% |
| Month 9 | 4.02964% | 5.84033% |
| Month 10 | 3.95818% | 5.76556% |
| Month 11 | 4.74161% | 5.2742% |
| Month 12 | 5.77242% | 4.08011% |
